# Accessibility and Disability Inclusion Among Top-Funded U.S. Undergraduate Institutions

**DOI:** 10.1101/2022.02.17.22271105

**Authors:** Jessica Campanile, Caroline Cerilli, Varshini Varadaraj, Fiona Sweeney, Jared Smith, Jiafeng Zhu, Gayane Yenokyan, Bonnielin K. Swenor

**Author notes:** **Corresponding author:**. Address: Johns Hopkins Disability Health Research Center, Johns Hopkins University, 525 N. Wolfe Street, Baltimore, MD, 21205, USA. **Data and materials availability:** Data is available on the Johns Hopkins Disability Health Research Center website at [https://disabilityhealth.jhu.edu/inclusiondashboard/].

## Abstract

**Background:** There is limited data to assess, track, or quantify accessibility and disability inclusion across universities.

**Objective:** This study assessed disability inclusion and accessibility at the top 50 National Institutes of Health (NIH)-funded undergraduate programs in the United States. We hypothesized that there is no association between NIH funding and the University Disability Inclusion Score

**Methods:** A novel tool, the University Disability Inclusion Score assessed disability inclusion and accessibility using 10 indicators spanning 4 categories: (1) accessibility of built and virtual environment, (2) public image of disability inclusion, (3) accommodations processes and procedures, and (4) grievance policy. Based upon the total points (out of a total score of 100), each university was assigned a letter grade (A-F).

**Results:** Of the top 50 NIH-funded institutions, 6% received an A grade on the Score, while 60% received D or F. The mean scores were 15.2 (SD=5) for accessibility of built and virtual environment (20 points), 10 (SD=3) for public image of disability inclusion (20 points), 30.6 (SD=10) for accommodations processes and procedures (50 points), and 8.1 (SD=3) for grievance policy (10 points).

**Conclusions:** Our findings suggest room for improvement in disability inclusion and accessibility among top university recipients of NIH funding. To provide an equitable academic experience, universities must prioritize disability inclusion.

## Introduction

People with disabilities are underrepresented in higher education. In fall 2019, 16.6 million students were enrolled in undergraduate degree-granting postsecondary programs.^1^ Approximately 19% of the undergraduate students have a disability^2^. But only about 1/3 of these students graduate within eight years.^3^ As higher education focuses on diversity and addressing inequities, it is imperative to identify and address barriers for students with disabilities and close these education gaps.

Increasingly, colleges and universities are taking a data-driven approach to develop policy and allocate resources. Yet there are critical data gaps that limit the development of evidence-based approaches to disability inclusion and accessibility across higher education. Data are needed to improve the inclusion and outcomes of people with disabilities in higher education.

The Rehabilitation Act of 1973 mandates that any institution receiving federal funding, such as grants from the National Institutes of Health (NIH), is required to ensure that people with disabilities do not face exclusion or discrimination from its programs and activities.^4^ However, without data to track and identify gaps in accessibility, federal agencies, such as the NIH, cannot assess an institution’s compliance with these regulations when allocating funding.

We created a novel tool, the University Disability Inclusion Score, to evaluate disability inclusion and accessibility of U.S. undergraduate academic institutions. This project scored the top 50 NIH funded institutions and created the University Disability Dashboard to publicly share this information.^5^ Using this data, we tested our hypothesis that there is no association between NIH funding (in US dollars) and score on the University Disability Inclusion Score.

## Methods

### University Selection

NIH research funding data was obtained from NIH RePORT (https://report.nih.gov/award/score.cfm) on June 21, 2021. This resource ranks aggregate NIH funding across universities for each fiscal year. University data from fiscal year 2020 was used and limited to domestic higher education institutions. Medical, dental, or veterinary institutions, as well as universities offering exclusively graduate degrees, were excluded. Our sample included the top 50 undergraduate degree granting U.S. institutions.

Institutional Review Board approval was not necessary as these data are publicly available and does not constitute human subject research.

### Coding Process

Two primary coders, J.C. and C.C., both disabled individuals with experience as undergraduate students, abstracted data for the 50 universities synchronously. The initial phase was an iterative process from June-September 2021 where these primary coders reviewed each university’s publicly available webpages together for a preliminary sample of 10 universities. The primary coders discussed the indicators with B.S. and updated the scoring protocol, point allocations, and key definitions based on data availability. Upon agreeing on an updated protocol, the primary coders coded again any indicators that were changed in the iteration process for all coded universities. This process was completed 5 times, once each at the completion of 10, 20, 30, 40, and 50 universities.

Two secondary coders, F.S. or V.V. independently abstracted and scored data for all 50 universities, scoring 26 and 24 universities, respectively. Upon completion of all coding, the primary and secondary coders held adjudication meetings to compare and resolve any discrepancies in the scoring.

All coding was completed in Microsoft Excel by J.C., who designed a custom instrument in Excel that allowed coders to input the description of a university’s indicator-specific performance, and subsequently calculated the corresponding numerical point allotment. The data collected excluded any informational videos or other multimedia presentations on the websites accessed. Links to outside sources, such as accessibility services initiatives at other universities, or to the U.S. Department of Education website, were not included.

### Scoring

The University Disability Inclusion Score assigns values to each university across 10 indicators (**Table 1**), each worth a maximum of 10 points for a total of 100 possible points per university. To enhance usability of this data, these indicators were grouped into four categories based on the following themes:

**Table 1.** Summary of University Disability Inclusion Score categories, indicators, and point allocations

| Category | Indicator | Point Allocations |
| --- | --- | --- |
| Accessibility of Built and Virtual Environment | Accessibility of Built Environment | 10 points: Map includes 3 – 4 key features (accessible paths, restrooms, parking, entrances).<br>5 points: Map includes 1 – 2 key features.<br>0 points: Map includes 0 key features or no map located. |
| Accessibility of Built and Virtual Environment | Accessibility of Virtual Environment | 10 points: Web accessibility score $\geq 7$.<br>5 points: Web accessibility score $< 7$ and $\geq 4.3$.<br>0 points: Web accessibility score $< 4.3$. |
| Public Image of Disability Inclusion | Student Disability Services Office Statement | 10 points: Disability services office statement included encouraging language.<br>5 points: Statement included only legal language.<br>0 points: No statement located. |
| Public Image of Disability Inclusion | Statistics and Estimates of Disabled Students, Staff, and Faculty | 10 points: Data for disabled students, staff, and faculty published on a university website.<br>5 points: Data for disabled students only published on a university website.<br>0 points: No data located. |
| Accommodations Processes and Procedures | Clear Point or Route of Contact for Requesting Accommodations | 10 points: Accessible software or form, or multimodal (phone **and** email information) contact provided for accommodations applications.<br>5 points: Only phone **or** email contact provided.<br>0 points: No route of contact provided. |
| Accommodations Processes and Procedures | Confidentiality of Disability and Accommodations Status | 10 points: Statement that asserts confidentiality maintenance and defines responsible parties.<br>5 points: Statement that generally mentions confidentiality without detailing relevant parties and covered information.<br>0 points: No information about confidentiality located. |
| Accommodations Processes and Procedures | Student and Disability Services Office Rights and Responsibilities | 10 points: Rights and responsibilities of students and disability services office available on disability services office website.<br>5 points: Rights and responsibilities only listed for one party, or only one of responsibilities/rights listed for both parties.<br>0 points: Rights and responsibilities information incomplete or not located. |
| Accommodations Processes and Procedures | Faculty Rights and Responsibilities | 10 points: Rights and responsibilities of faculty available on disability services office website.<br>5 points: Only rights or responsibilities listed.<br>0 points: Rights and responsibilities information not |
|  |  | located. |
| Accommodations Processes and Procedures | Timeline for Accommodations Application and Approval | 10 points: Flexible timeline provided with clear expectations for application and approval/rejection timing.<br>5 points: Static deadline provided.<br>0 points: Timeline information not located. |
| Grievance Policy | Existence and Handling of Grievance Policy | 10 points: Grievance policy exists, is listed or linked on disability services office webpage, and is handled by a third party.<br>5 points: Grievance policy exists and is handled by a third party, but no information exists on the disability services office webpage.<br>0 points: Grievance policy exists and is handled internally by the disability services office, or no grievance policy information located. |

1. accessibility of built and virtual environment (2 indicators, 20 points)
2. public image of disability inclusion (2 indicators, 20 points)
3. accommodations processes and procedures (5 indicators, 50 points)
4. grievance policy (1 indicator, 10 points)

For each indicator, scoring options included 10, 5, or 0 points based on the criteria outlined below. All requirements for a score must be met to attain that score.

#### Development of Scoring Indicators

To inform this work, we performed a literature review of relevant existing disability policies and considerations in higher education in the U.S. Our scoring criteria and indicators were developed based on prior work examining the presence of disability policies at graduate medical institutions and their compliance with policy requirements.^6^

### Accessibility of Built and Virtual Environment

#### Accessibility of Built Environment

Only publicly available data on university websites were used, and this indicator did not score the level of accessibility of a given university’s campus. It instead indicates whether a university made the effort to create and publish a map with clearly marked accessibility features. Coders assessed publicly available campus maps for the marking of four features: accessible (1) paths, (2) restrooms, (3) parking, and (4) entrances. Assessments for paths included grade of slope, or elevator or stair presence. Coders conducted an internet search for “[UNIVERSITY NAME] campus map,” and if that map did not include accessibility information, searched “[UNIVERSITY NAME] accessible campus map.” A university received 10 points if a publicly available campus map included 3-4 of the features, 5 points if the map included 1-2 of the features, and 0 points if the map included none of the features or no map could be located.

#### Accessibility of Virtual Environment

For each university, a large sample of web pages were collected and analyzed by WebAIM at Utah State University using the WAVE web accessibility testing tool. WAVE is a commonly utilized testing tool and a free, online version of WAVE is publicly available at https://wave.webaim.org/.^7^ The university’s main web site, undergraduate admissions information web site, and student disability services office (DSO) web site were analyzed. Up to 500 web pages for each of these three site areas were tested using automated testing processes. Each university’s site was assigned an automated accessibility score based on the number of detected errors, the density of errors (number of errors by number of web page elements), and number of likely or potential accessibility errors. This score was determined based on alignment to the WebAIM Million^8^ sample of one million homepages to indicate how the tested university web site compared to web pages generally across the web. Expert testers then evaluated the university’s main homepage, undergraduate admissions information page, DSO page, and one randomly selected page from the sample above based on 10 aspects of web accessibility: (1) accuracy of the document’s defined language, (2) appropriateness of image alternative text, (3) impact of empty links and buttons, (4) impact of labeled or unlabeled form inputs, (5) low contrast content, (6) appropriateness of page title, (7) animation and movement, (8) keyboard focus indicators, (9) keyboard accessibility, and (10) page reflow/responsiveness. Scores based on potential impact of these accessibility issues on end users with disabilities, as well as an overall score for page accessibility, were provided for each web page. The manual and automatic scores were then averaged to generate an overall accessibility score. A university received 10 points if its total score was ≥7, 5 points if its total score was ≥4.3 and <7, and 0 points if its total score was <4.3.

### Public Image of Disability Inclusion

#### Student Disability Services Office Statement

Coders analyzed the publicly available DSO mission and vision statements, and where not available, the relevant sections describing the office’s role on the “About” or “Home” webpages on the DSO website. Statements were given a score based upon the presence of encouraging or supportive language, exclusively legal requirements, or lack of statement altogether. Encouraging or supportive language included: valuing disability as a form of diversity, welcoming disabled students to the university, encouraging disabled students to apply for accommodations, and stating a commitment to moving beyond what is legally required to alter university culture and educate the community. A commitment to “full access” to education for disabled students was not considered sufficiently encouraging or supportive, as this is required by law. A university received 10 points if a publicly available statement on the DSO website included encouraging or supportive language, 5 points if the statement included exclusively legal language, and 0 points if coders could not locate such a statement.

#### Statistics and Estimates of Disabled Students, Staff, and Faculty

To ascertain the presence of disability data on publicly accessible university websites, coders conducted an internet search using the following search terms: “[UNIVERSITY NAME] disability statistics” and “[UNIVERSITY NAME] number of disabled students, staff, and faculty.” Data had to exist on a main university webpage, including but not limited to webpages from the departments of admissions, public relations, diversity and inclusion, or disability services to meet criteria. Disability data found solely in institutional research documents, student or professional journalism, or other outside sources were not included. A university received 10 points if disabled student, staff, and faculty statistics or estimates were publicly available on a DSO website, admissions website, or with other protected class and diversity information, 5 points if only data on disabled students were published in one of the aforementioned locations, and 0 points if no disabled student, staff, and faculty data were found on any publicly facing website.

### Accommodations Processes and Procedures

#### Clear Route of Contact for Requesting Accommodations

Coders assessed the route students could use to request accommodations at each university. Information was collected regarding the presence of accommodations request software and forms, provision of multi-modal (both phone and email) contact information for a DSO professional, or a lack of clear route for students requesting accommodations. To maximize flexibility and accessibility of options, a university received 10 points if the DSO employed an accessible accommodations request software or form such as a PDF or listed both phone and email contact information for relevant DSO employees. A university received 5 points if only either phone or email contact information was listed, and 0 points if the DSO offered no clear point or route of contact for requesting accommodations.

#### Confidentiality of Disability and Accommodations Status

Coders searched for statements from DSO websites regarding the confidentiality of a student’s records. Ideal statements included information on who maintains the confidential records, if such information would appear on a student’s academic record, how information is shared about a student’s disability and accommodations, and how a student may grant others (such as parents) access to these records. A university received 10 points if the DSO website clearly stated that confidentiality of a student’s disability and accommodations status would be maintained and by whom. Statements had to include that the DSO maintains confidentiality to receive full points, as only mentioning faculty responsibility to maintain confidentiality was deemed insufficient. Universities were given 5 points if the DSO website included a general mention of confidentiality with no clear details regarding responsible parties or covered information, and 0 points if the website included no mention of confidentiality.

#### Student and Disability Services Office Rights and Responsibilities

The presence of rights and responsibilities for both students and the DSO were examined. Clearly labeled student rights include (but are not limited to) the right to confidential records, to request accommodations, and to file a grievance if they believe they have been treated unfairly. Student responsibilities include the responsibility to apply for accommodations from the DSO, and to communicate with faculty. DSO rights include the right to deny a student’s accommodation request if they determine the request is unreasonable. DSO responsibilities include the responsibility to maintain a student’s confidential records and to provide services afforded to students under applicable laws, such as the Americans with Disabilities Act or Section 504 of the Rehabilitation Act. Rights and responsibilities must be centrally located in an exhaustive list. Mere suggestions, as opposed to clear requirements, that a student or office should adhere to these rights and responsibilities were viewed as insufficient and subsequently not scored. A university received 10 points if the rights and responsibilities of both disabled students and the DSO were made clear on the DSO website, 5 points if the rights and responsibilities of only disabled students or the DSO, or only rights or responsibilities for both parties were made clear, and 0 points if no rights or responsibilities were published for either party, or if this information was otherwise incomplete.

#### Faculty Rights and Responsibilities

The rights and responsibilities of faculty were also examined. Clearly outlined faculty rights include information about the right to raise concerns to the DSO if they deem an accommodation to fundamentally alter essential components of their course. Faculty responsibilities include the responsibility to implement all reasonable accommodations and to maintain confidentiality of a student’s disability status or diagnosis. Rights and responsibilities must be centrally located in an exhaustive list. Suggestions, as opposed to requirements, that faculty should adhere to these rights and responsibilities were viewed as insufficient. A university received 10 points if the rights and responsibilities of faculty regarding accommodations and disabled students were made clear on the DSO website, 5 points if only either the rights or responsibilities of faculty were published, and 0 points if the DSO website did not detail the rights and responsibilities of faculty.

#### Timeline for Accommodations Application Approval

Coders examined the DSO webpage for a clear timeline from when a student requested accommodations to when this request would be approved. Different types of accommodations can take varying times to be implemented. However, it is important to provide students with expectations of accommodations approval or rejection timelines for the purposes of registering for and choosing courses, as well as planning medical appointments to obtain required disability documentation. A university received 10 points if the DSO website offered a flexible timeline (ex. 3 weeks) for the time between an accommodation request submission and approval or rejection, 5 points if the site offered a static deadline (ex. by August 1^st^), and 0 points if the site gave no clear timeline for student request and office approval or rejection of accommodations.

### Grievance Policy

#### Existence and Handling of Grievance Policy

Coders searched for a clear policy that describes how students may file a complaint about discrimination relating to disability on the DSO website. Coders investigated if the DSO handled these issues, or if the issue could be raised to a third party, defined here as an individual outside of the immediate DSO, such as an office for institutional equity and diversity or an ombudsman. If coders were unable to locate the grievance policy, they conducted an internet search for “[UNIVERSITY NAME] disability grievance policy.” Appeals processes that apply only to the accommodation request and approval process and not discrimination were not considered to be a full grievance policy. Universities must list a disability-specific non-discrimination policy for students, as a general non-discrimination policy is insufficient for disability-specific concerns, and disabled employees and students are protected by different legal structures. A university received 10 points if a student disability discrimination grievance policy existed, was handled by a third party, and was clearly listed or linked on the DSO website, 5 points if such a policy existed and was handled by a third party, but no relevant information was present on the DSO website, and 0 points if a policy existed and was handled internally by the DSO, or if no information whatsoever could be found regarding the policy on any publicly facing university website.

### Statistical Analyses

Based upon the total points (out of a total score of 100), each university was assigned a letter grade (A-F) using the following categorization: 90-100 points received an A, 80-89 points received a B, 70-79 points received a C, 60-69 points received a D, and 0-59 points received an F. The mean, standard deviation, median, range, and interquartile range (IQR) were calculated for the overall score and scores for each of the subcategories. Counts of institutions receiving each overall grade and point allotment for each of the four scored categories were plotted as bar graphs. The number of times (frequency and percentage) a university’s indicator score changed during adjudication was also calculated. A simple linear regression model was used to examine the association between NIH funding (in million dollars) and a university’s score. The university score was the outcome and the NIH funding was a covariate. Statistical analyses were completed using R statistical software (version 4.1.1).

## Results

Among the 50 top NIH-funded undergraduate institutions, the majority received a grade of D or F (≤69 points out of 100 possible points), (**Figure 1**). The mean overall score across the sample was 64 (SD=14), and the median score was 60 (IQR=15, range=30–90). During adjudication, of the 450 indicators that were scored in total (50 schools x 9 indicators; not including the WebAIM scoring), there were 51 (11.3%) indicators where primary and secondary coders disagreed, and changes were made to the point allocations.

**Figure 1.**
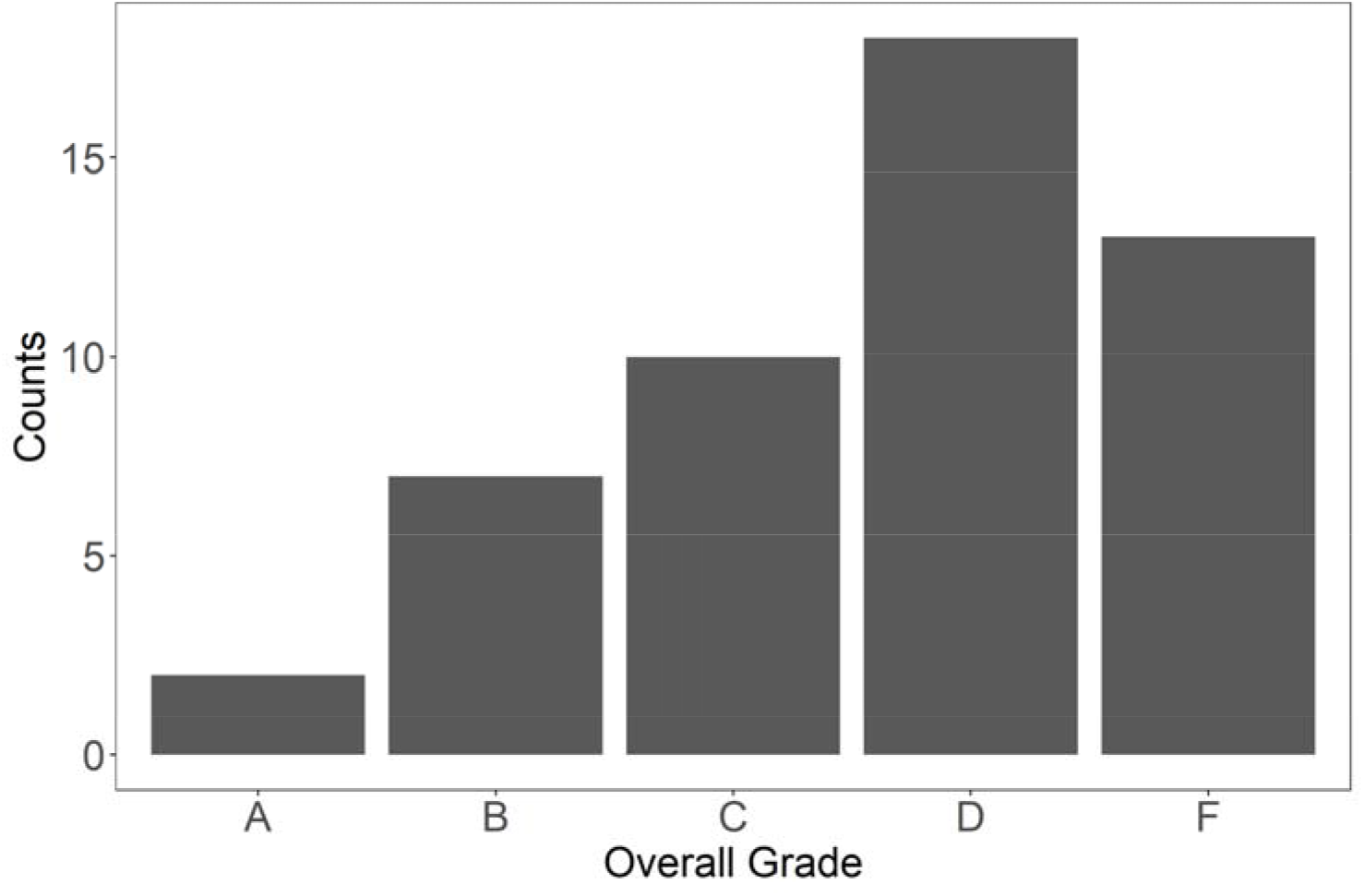
Count of institutions receiving each overall grade

The accessibility of built and virtual environment (20 points) mean score was 15.2 (SD=5) and the median score was 15 (IQR=8.75). Of the two indicators included in this category, universities performed better in accessibility of the virtual environment (mean=8.9/10 points) than accessibility of the built environment (mean=6.3/10 points), (**Figure 2**). The public image of disability inclusion (20 points) mean score was 10 (SD=3) and the median score was 10 (IQR=0). Universities performed better in student disability services office statement (mean: 8.8/10) than in statistics on disabled students, staff, and faculty (mean=1.2/10). The accommodations processes and procedures (50 points) mean score was 30.6 (SD=10) and the median score was 30 (IQR=17.5). Of the five indicators included in this category, universities performed best in clear point or route of contact for requesting accommodations (mean: 9.7/10) and worst in faculty rights and responsibilities (mean=3.9/10). grievance policy (10 points) included a single indicator (existence and handling of grievance policy), received a mean score of 8.1 (SD=3) and a median score of 10 (IQR=3.75).

**Figure 2.**
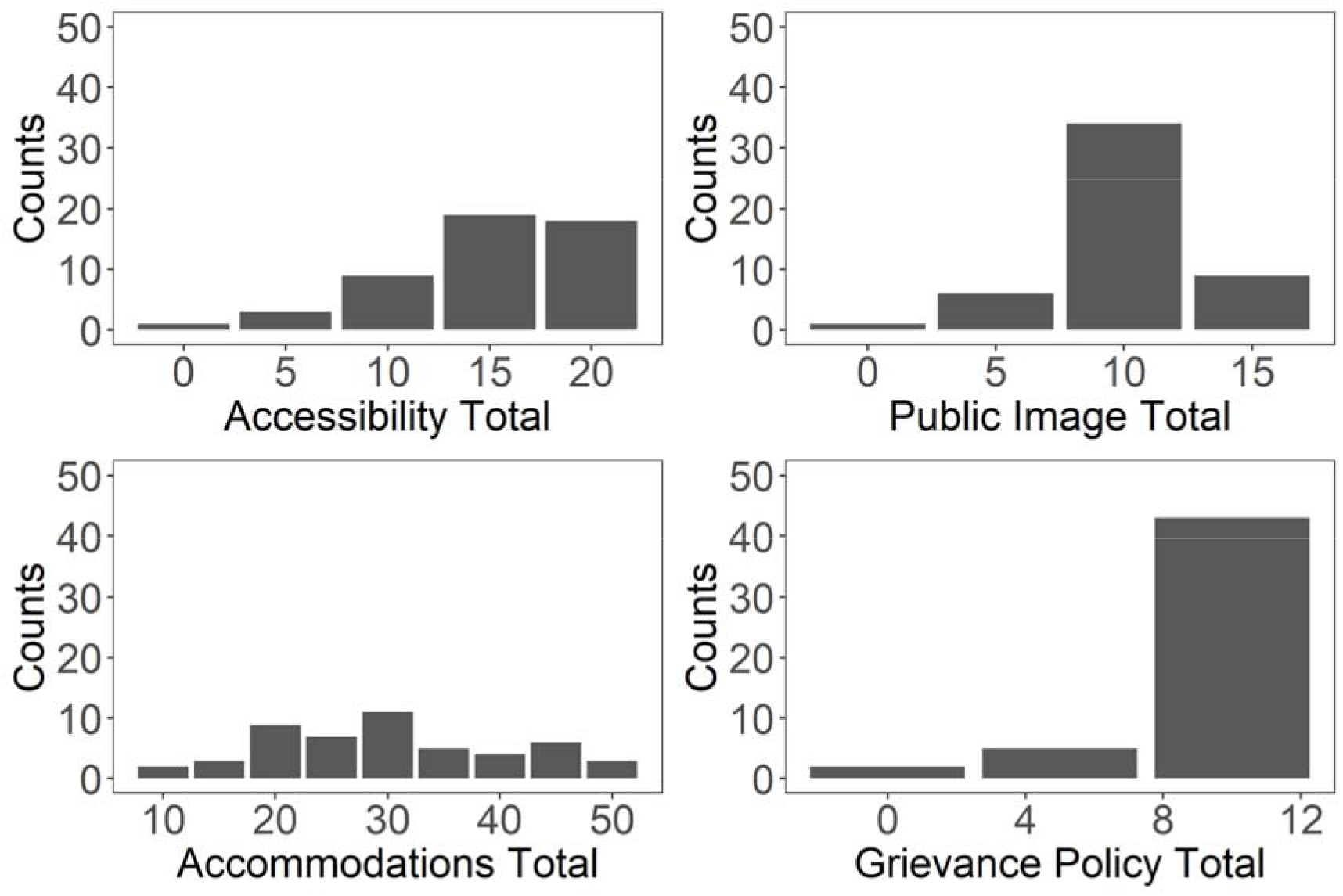
Count of institutions receiving each point allotment for the four scored categories.

Across the four categories, universities earned the highest percentage of points (81%) in grievance policy, and the lowest percentage of points (50%) in public image of disability inclusion (**Figure 3**).

**Figure 3.**
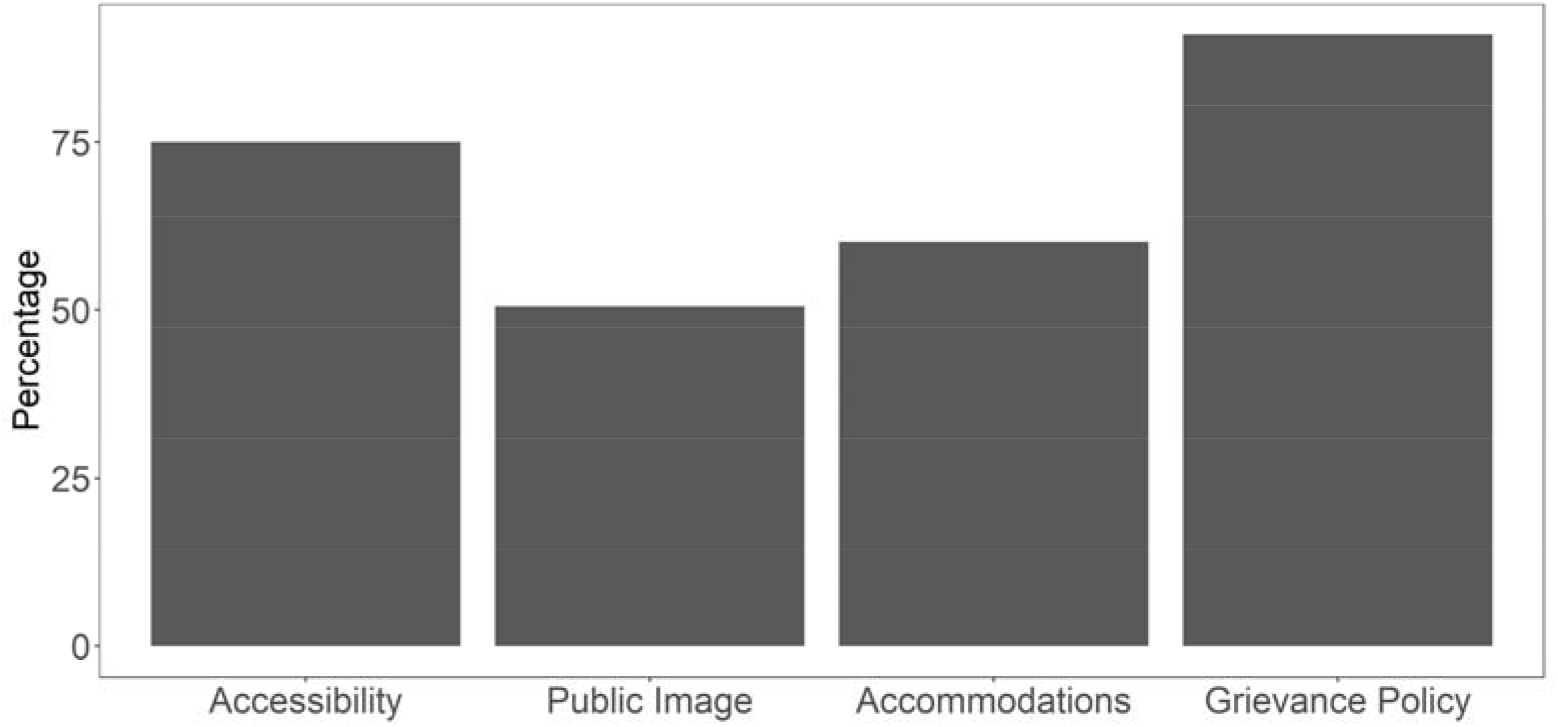
Percentage of points earned by institutions in each of the four scored categories.

In regression analysis, no association was noted between NIH funding and a university’s score (ß=0.009, 95% CI= -0.011, 0.029, p-value=0.379).

## Discussion

Our results indicate that the majority of the top 50 NIH-funded undergraduate universities have significant room for improvement on disability inclusion and accessibility, as 60% received a score of D or F on the University Disability Inclusion Score. Low scores indicate a possible lack of adherence to the Americans with Disabilities Act (ADA) and Section 504 of the Rehabilitation Act and suggest that many universities may need to take action to rectify their deficiencies in these areas.

In 2011-2012, 25% of first-year disabled undergraduates, 35% of second-year disabled undergraduates, and 36% of third-year disabled undergraduates left their academic institutions and did not return.^9^ To increase representation of disabled individuals in professions requiring graduate schooling as well as undergraduate degree completion, such as many professions in science, technology, engineering, mathematics, and medicine (STEMM), it is vital to pinpoint the accessibility barriers that make the application and graduation processes disproportionately difficult for disabled students. This project established a method to compare disability inclusion and accessibility across universities, which is a first step to developing data-driven approaches to closing gaps in retention and graduation rates for disabled students.

Of the four categories scored in this study, universities performed best in the Grievance Policy category, likely due to the clear guidance outlined in Section 504 of the Rehabilitation Act regarding grievance procedures for disability discrimination. Universities performed worst in the public image of disability inclusion category as the statistics on disabled students, staff, and faculty indicator was the lowest scoring overall, with a mean of 1.2 out of 10 points. This indicates the need for universities to actively collect data on disability from their students and employees in the same manner that they do for other protected classes such as gender and race. The clear point or route of contact for requesting accommodations indicator was the highest scoring overall, with a mean of 9.7 out of 10 points. In higher education, as opposed to elementary or secondary school, students must request their own accommodations. This process can include feelings of discomfort, apprehension, and fear of stigma, leading students to be less likely to request accommodations.^10^ As such, a clear route for requesting accommodations is integral to mitigating these barriers and encouraging student requests.

Overall, universities received low average scores for the student and disability services office rights and responsibilities, and faculty rights and responsibilities indicators, 4.3 and 3.9 out of 10 respectively. This suggests room for improvement in communicating the roles of relevant parties in accommodations request and provision. Universities received an average score of 4.4 out of 10 points for timeline for accommodations application and approval, indicating a need for setting clearer expectations for student accommodation application and receipt timelines.

Federal funding bodies can support improved accessibility and inclusion for individuals with disabilities by prioritizing these aspects of an institution in their funding decisions. As the Rehabilitation Act of 1973 outlines, any institution receiving federal funding is prohibited from excluding or discriminating against disabled people, and these results suggest federal funding agencies are not leveraging their role to advance disability inclusion in these settings.

## Limitations

The strengths of this study include a systematic approach to developing a quantitative score that targets important aspects of disability inclusion and clearly defined scoring guidelines and adjudication processes. However, the limitations should be considered. This data was coded over a period of 4 months, and information on university websites may have changed during or after the coding process. Estimates of disability prevalence provided by DSOs may not be fully representative. Such data are largely dependent on accommodation disbursement, which is not an accurate representation of students who may identify as disabled for reasons such as limited access to healthcare or diagnostic tests, or stigma associated with receiving accommodations. Additionally, some universities provided detailed statistics while others provided general estimates of prevalence. Due to a dearth of data, both statistics and estimates fulfilled the criteria for this study. Ideally, universities should collect and publish statistics that can be compared across institutions.

This study is limited to publicly available data listed on university websites. There may be additional accessibility practices and disability inclusion resources on internal or password-protected webpages that were not included in these data. We also acknowledge that higher scores and the presence of inclusive language, disability statistics, and other inclusion indicators on a university website do not necessarily translate to an inclusive institutional environment for students with disabilities. An important next step may be to survey students at the campuses of these institutions and understand their lived experiences as compared to the University Disability Inclusion Score. This scoring is therefore not a definitive ranking of qualitative student experience at these institutions. However, there is no mechanism currently available to qualitatively assess on-the-ground disabled student experiences at every university that receives federal funding. This study serves to lay the groundwork for tracking accessibility and disability inclusion data at universities nationwide, and to signal to institutions that receive federal funding that, even with a scoring system that is not all-encompassing, they are failing to reach an appropriate standard.

## Conclusions

Using a novel tool, the University Disability Inclusion Score, we found that the majority of the top 50 NIH-funded universities have considerable room for improvement on disability inclusion and accessibility, as 60% received a score of D or F. The road to an A – and closing gaps to higher education for people with disabilities – is achievable. It is time for universities to prioritize disability inclusion and accessibility and for federal funding agencies to consider these efforts as part of grant review criteria.

## Data Availability

All data produced are available online at https://disabilityhealth.jhu.edu/inclusiondashboard/

## Acknowledgements

The authors have no funding or conflicts of interest to disclose.

